# Immunotherapy Efficacy Prediction in Cancer: An Artificial Intelligence Approach with Unannotated H&E Whole-Slide Images

**DOI:** 10.1101/2024.02.05.24301493

**Authors:** Gabriel Domínguez Conde, Talha Qaiser, Evan Wu, Carlos Eduardo de Andrea, Jennifer Shields, Ronen Artzi, Harish RaviPrakash, Kenneth Irabor, Paul Metcalfe, Joachim Reischl

**Affiliations:** Oncology R&D, AstraZeneca, Cambridge, UK; CPSS, R&D, AstraZeneca, Cambridge, UK; Department of Pathology, Clínica Universidad de Navarra, Pamplona, Spain

## Abstract

Developing a solution to predict clinical outcomes for immunotherapy that is accurate, scalable, affordable, clinically meaningful, and globally accessible is an unmet medical need. Precise prediction of patient response to immunotherapy from pretreatment biopsy images will enable the delivery of immuno-oncology drugs to suitable patients and reduce the risk of administering unnecessary toxicity to patients who do not benefit from it. We propose an AI-based framework to produce stratifying algorithms that only need routinely obtained unannotated hematoxylin and eosin (H&E)-stained whole slide images. This design choice eliminates the need for pathologist annotations, ensuring affordability and scalability. Our solution, developed with data from 418 durvalumab patients, was validated both for head and neck squamous cell carcinoma and non-small cell lung cancer with data from 283 durvalumab patients, demonstrating its versatility and ease of adaptation for different indications. The results obtained using test data from clinical trials, different from training data, exhibit clinically meaningful improvement between those classified as positive and negative. For median overall survival (OS), the enhancement is in the range [55.9%, 198%] and [0.49, 0.70] for the hazard ratio for OS. For median progression-free survival (PFS), the improvement ranges within [39%, 195%], while the hazard ratio is within [0.46, 0.86] for PFS. Our solution complements the current biomarker, programmed death lig– and 1, for immunotherapy therapy, presenting an opportunity to develop more accurate solutions. In addition, as the algorithm was developed in a hypothesis-free approach, the analysis of the converged solution may enhance expert understanding of the pathomechanisms driving the response to immunotherapy. Its scalability and accuracy, combined with the AI-based engineering nature of the solution, bring the opportunity of being globally deployed using the cloud. Our technique has the potential to unlock opportunities not available for patients before by enabling the generation of efficient and affordable diagnoses for immunotherapy.

## 1 Introduction

Lung cancer and Head and Neck cancer together account for over 20% of cancer deaths worldwide [1, 2]. Recent advances in immune checkpoint inhibitors (ICI) have led to its widespread use as a therapeutic strategy across multiple tumor types. In particular, responses to antibodies inhibiting programmed death 1 (PD-1)/programmed death ligand 1 (PD-L1) in advanced non-small cell lung cancer (NSCLC) and head and neck squamous cell carcinoma (HNSCC) have demonstrated robust and durable anti-tumor activity leading to its incorporation as first-line treatment options for newly diagnosed patients with these cancers in the recurrent or metastatic settings [3–10]. Nonetheless, despite the repeated demonstration of the superiority of ICI over traditional chemotherapy, only a small subset of NSCLC and HNSCC patients respond to ICI [11, 12].

As such, there has been a growing interest in identifying predictors of benefit to ICI. In the present day, the selection of patients for PD-(L)1 inhibitors in HNSCC and NSCLC primarily relies upon detecting the expression of PD-L1 by immunohistochemistry (IHC) staining [13]. These assays use a variety of detection platforms at different levels (protein, mRNA), employ diverse biopsy and surgical samples, and have disparate positivity cut-off points and scoring systems, all of which complicate the standardization of clinical decision-making [14]. In addition, inconsistencies in using PD-L1 expression as a biomarker for response to immunotherapy have emerged: many patients with high PD-L1 expression do not respond to ICI [15–18], and patients with no discernible PD-L1 expression may show remarkable responses to ICI. PD-L1 expression status can vary significantly based on the assay used (i.e., the validated PD-L1 IHC assays are Ventana SP142, Ventana SP263, and Dako 22C3), and therefore may vary across medical institutions [19]. As the evidence for immunotherapy continues to expand, a critical and unmet clinical need is the identification of reliable assays to predict response to ICI. In order to address these outstanding clinical needs, researchers have focused on opportunities brought by combinin two technological breakthroughs: digital pathology and the revolution of computer vision brought through the wide use of deep learning (DL).

Digital pathology’s emergence as a scientific discipline directly results from the digitizing of traditional practices. In essence, high-resolution digital scanners capture detailed whole-slide images (WSIs) from glass slides. Adoption grew when the U.S. Food and Drug Administration (FDA) gave clearance to the first WSI system in 2017 [20], updating the traditional workflow of the pathologist by facilitating content sharing as well as catalyzing the adoption of new digital techniques to assist pathologists in working with more efficiency and accuracy. Recently, multiple studies compared the histopathology diagnostic performance between digital pathology WSIs and glass slides under the microscope [21]. The clinicians observed similar diagnostic performance and reported error rates. Various AI-driven applications in digital pathology emphasize reducing the turnaround time for pathologists.

Furthermore, the performance of the state-of-the-art algorithms of traditional computer vision applications has experienced a boost obtained by the extensive adoption of artificial intelligence (AI) in general and DL in particular [22], driven by its success [23] in the general purpose classification dataset ImageNet, a large dataset used to benchmark the advances in image classification algorithms. DL in computer vision has demonstrated the ability to extract and learn features from large volumes of data to solve multiple tasks, including image classification, object detection, semantic segmentation, and face recognition. The applications of deep learning in the field of medical imaging have increased significantly [24], including X-ray [25, 26], computed tomography (CT) [27, 28], magnetic resonance imaging (MRI) [29], positron emission tomography (PET) [30], and dermoscopy images [31].

The use of DL techniques in digital pathology has unique challenges when compared to other application domains (e.g., retail or manufacturing). These challenges come from two key aspects. Firstly, computing limitations due to the size of the WSIs often exceed the memory capacity of current graphics processing units (GPUs); a WSI can be gigabytes in size. Secondly, annotating WSIs is laborious and expensive, requiring pathologists to annotate, label, and classify regions of interest, a necessary step for training algorithms in the supervised setting.

Despite these limitations, the applications of DL in digital pathology are growing. For example, DL algorithms, developed as part of the CAMELYON challenge, were used for lymph node metastase detection in breast cancer [32] and found to have potential for pathological diagnosis. In prostate cancer, Bulten et al. developed a solution for automated tumor detection and segmentation from H&E stained slides [33]. Arvaniti et al. designed an approach to automatically grade cancer cell abnormality (Gleason grading) [34] in the prostate. More recently, the FDA authorized the first AI-based software [35], Paige Prostate, to assist pathologists by identifying the area of interest containing cancer cells on the prostate biopsy images. By using this solution, the pathologist can review those regions and help to make the diagnosis. Another application of deep learning in digital pathology is the prediction of genetic mutation. For example, in [36], the authors used convolutional neural networks (CNNs) to determine if the SPOP gene was mutated in prostate cancer. In the immuno-oncology (IO) therapy setting, Bains et al. developed an algorithm for automatic recurrence and risk prediction in NSCLC. Kapil et al. addressed the task of automated objective scoring of PD-L1 expression to identify patients who may respond to anti-PD-1/PD-L1 treatments [37].

DL in digital pathology for companion diagnostic (CDx) is a new area, as there is no cleared or approved device [38]. Unlike other approaches in this field, we propose to address the challenging task of predicting patient response to immunotherapy from pretreatment biopsy images. The successful stratification of patients into responders and non-responders will help oncologists to better prescribe treatment to patients. Towards this, we propose using weakly-supervised deep learning algorithms to stratify patients into two groups: responders to immunotherapy and non-responders. We named this approach artificial intelligence for predicting clinical outcome (AIPCO). Our hypothesis is that normal H&E-stained WSIs of patients contain encoded information regarding the responsiveness of the patients to immunotherapy.

To validate this hypothesis, we used five durvalumab trials, of which two are NSCLC trials (MYSTIC [39] and ARCTIC [40]), while the other three correspond to HNSCC (EAGLE [41], HAWK [42], and CONDOR [43]). Contrary to most of the existing approaches in the area that focus on supervised learning [44], we did not employ pathologist annotations but formulated the task as a clinical outcome-driven and hypothesis-free solution, as illustrated in the diagram shown in Fig. 1. The proposed approach involves training a tumor classification model followed by a response prediction at the patch level. Some of the benefits offered by this technique include a faster development cycle and enhanced scalability, as the pathologist’s time-consuming annotations are not required. In addition, our annotation-free technique may lead to novel discoveries on the relationship between WSI architectural features and clinical outcomes that human experts may not know. This information may be used to improve the accuracy of the prognostication of the disease.

**Figure 1:**
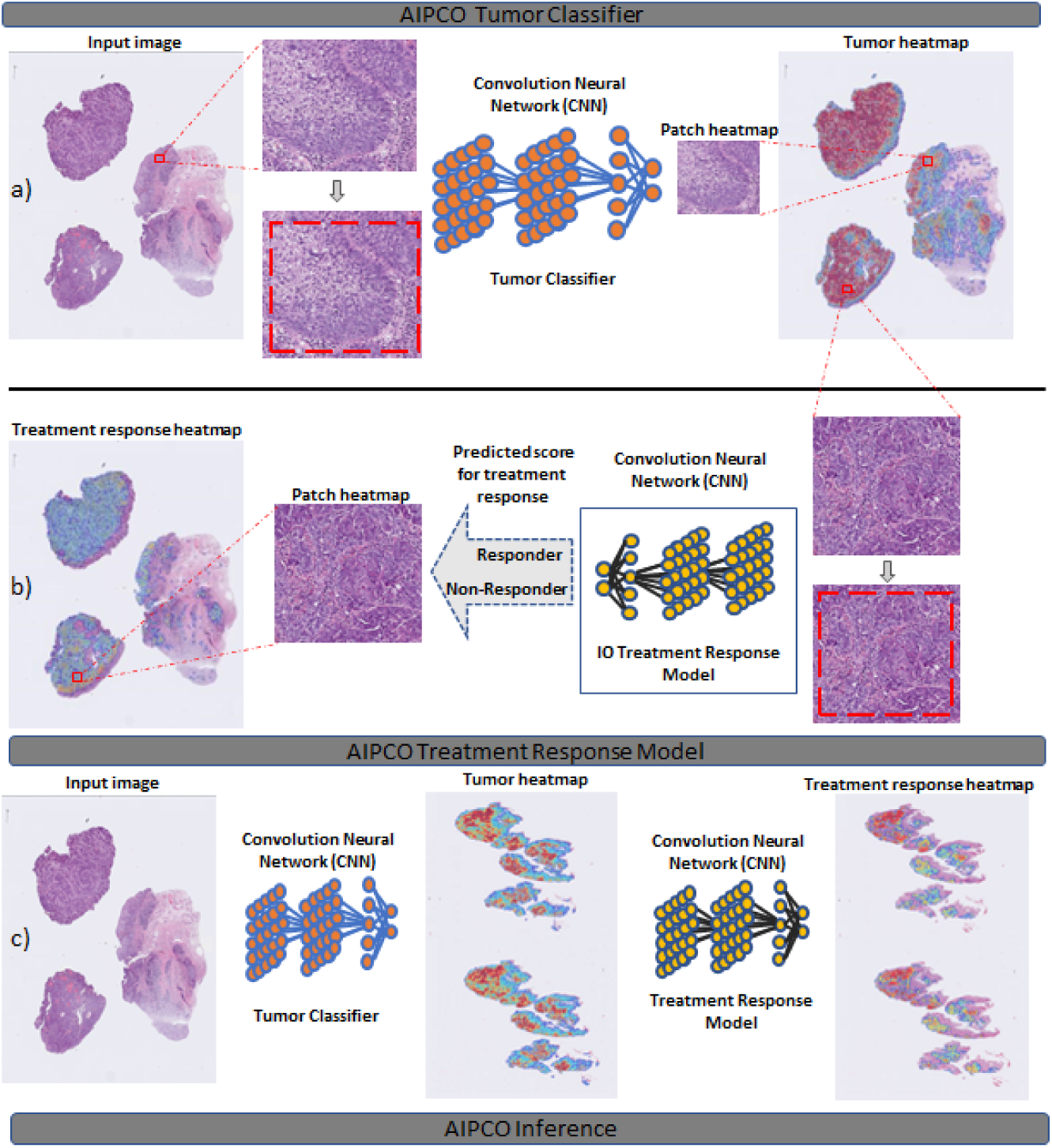
Overview of the proposed AIPCO treatment response pipeline for analyzing histology H&E stained WSI. On the top row a), the diagram shows the tumor classification method; for a given WSI, multiple patches were randomly selected. Different data transformations were applied to each patch during the training of the model. The output heatmap was then generated by aggregating patch-level predictions. In the middle row b), the flowchart represents the AIPCO treatment response pipeline. For each tumor predicted, a series of data transformations were applied before analyzing it to predict the likelihood of treatment response. A knowledge distillation model was used to learn discriminative features. On the bottom row c), the AIPCO inference strategy is shown; while testing, we run inference on the WSI level to perform tumor classification followed by prediction of treatment response.

## 2 Results

### 2.1 AIPCO trained with unannotated H&E whole-slide images predicts immuno-oncology therapy efficacy for non-small cell lung cancer and head and neck squamous cell carcinoma

#### Datasets

For the development of our framework AIPCO, we use data of patients who received durvalumab as part of Phase II or Phase III global multicenter clinical trials, as shown in Table 2. We use both the H&E WSIs and the associated clinical information of the patients, including the clinical outcome and the baseline characteristics. Since these modalities are needed, we can only use patients for which we have both. Due to the limited availability of patients with accessible H&E-stained WSIs compared to the total number of patients in the original studies, the cardinality of the dataset for developing our framework is reduced. To show the validity of our approach across indications, on the one hand, we use data from 292 HNSCC patients from EAGLE (n= 123), HAWK (n= 109), and CONDOR (n= 60). On the other hand, for NSCLC, the data used was obtained from 458 patients from MYSTIC (n= 312) and ARCTIC (n=146) clinical trials.

Our approach aims not only to assess the performance using subjects who were not used for training but also guarantee that these patients correspond to a clinical trial that is different from the one utilized for training. In this way, we validate the performance within the same study (held-out test set) and test it across other studies (inter-study validation). This is key to assessing the readiness of our solution for clinical practice. In addition, as training typically requires more data and we do not want to train biased algorithms, we used, as a training dataset, phase 3 clinical trials with patients with a variety of PD-L1 status. Aligned with these requirements, MYSTIC is the dataset used for training, and ARCTIC is the dataset used for testing for NSCLC. For HNSCC, EAGLE is selected for training, while HAWK and CONDOR are utilized for testing.

Using the Kaplan-Meier method [45], we graphically displayed PFS and OS in order to compare AIPCO responders AIPCO+ and AIPCO non-responders AIPCO-both for NSCLC and HNSCC.

#### AIPCO for head and neck squamous cell carcinoma

Fig. 2 for HAWK and Fig. 3 corresponding to CONDOR graphically show a significant improvement in PFS and OS for the AIPCO+ group for both outcomes, quantitatively expressed in Table 1, including objective response rate (ORR). In the case of HAWK, the median OS for the AIPCO+ responders group was 10.2 months as compared to 4.9 months for the AIPCO-group, 3.8 months vs. 1.9 months for the PFS case, and ORR is 25% for the AIPCO+ while for AIPCO-is 11%. Similarly, for CONDOR, mOS AIPCO+ patients was 14.9 months while for AIPCO-patients was 5.0 months, AIPCO+ patients showed an mPFS of 5.6 months vs. 1.9 months for AIPCO-, in terms of ORR the improvement goes from 5% for AIPCO-patients to 23% months for AIPCO+ subjects.

**Figure 2:**
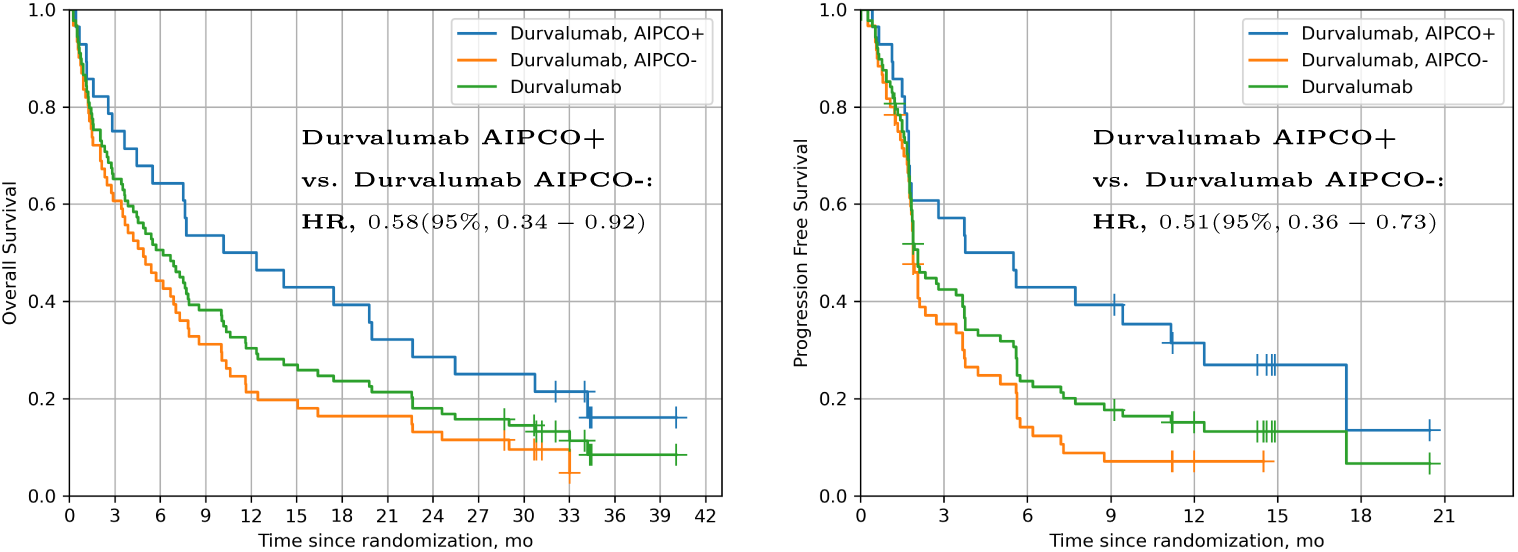
The overall survival (left) and progression-free survival (right) among HAWK patients who received durvalumab with status AIPCO+ (blue line) vs. AIPCO-(orange line). The AIPCO stratification is based on the AIPCO algorithm trained with EAGLE data. Green lines correspond to the curves of the HAWK patients before stratification using AIPCO. Both figures show the hazard ratio (HR) and the 95% confidence interval (CI) for comparing AIPCO+ and AIPCO-patients.

**Figure 3:**
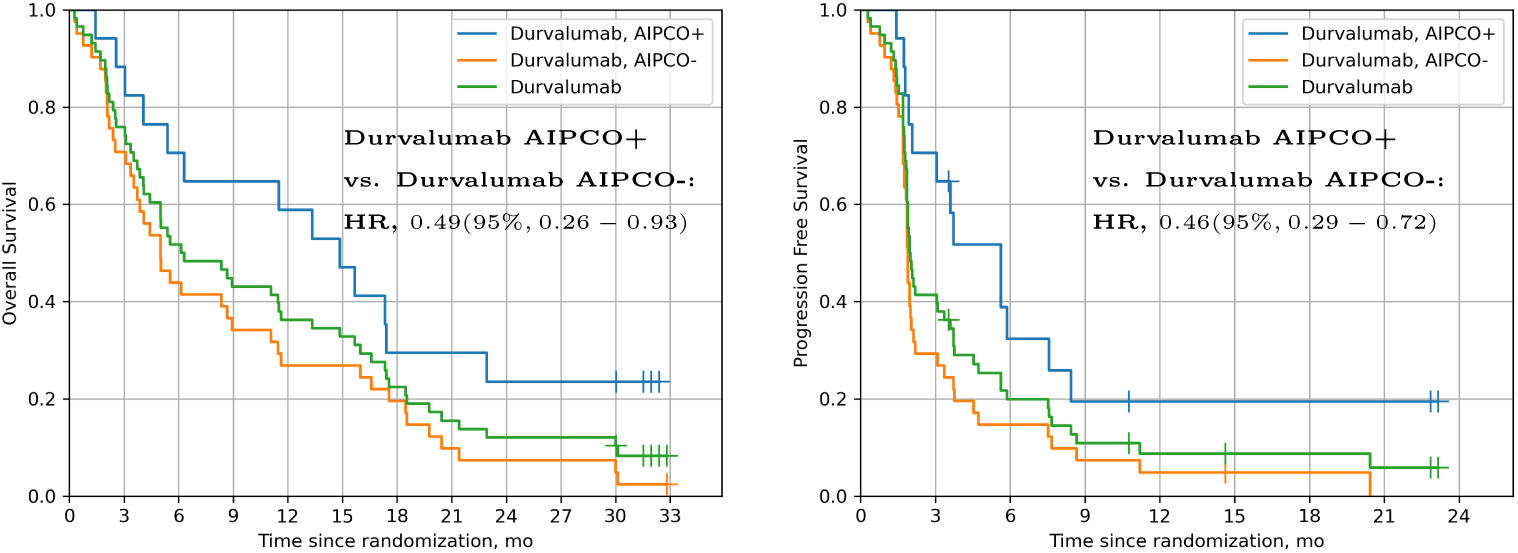
The overall survival (left) and progression-free survival (right) among CONDOR patients who received durvalumab with status AIPCO+ (blue line) vs. AIPCO-(orange line). The AIPCO stratification is based on the AIPCO algorithm trained with EAGLE data. Green lines correspond to the curves of the CONDOR patients before stratification using AIPCO. Both figures show the hazard ratio (HR) and the 95% confidence interval (CI) for comparing AIPCO+ and AIPCO-patients.

**Table 1:**
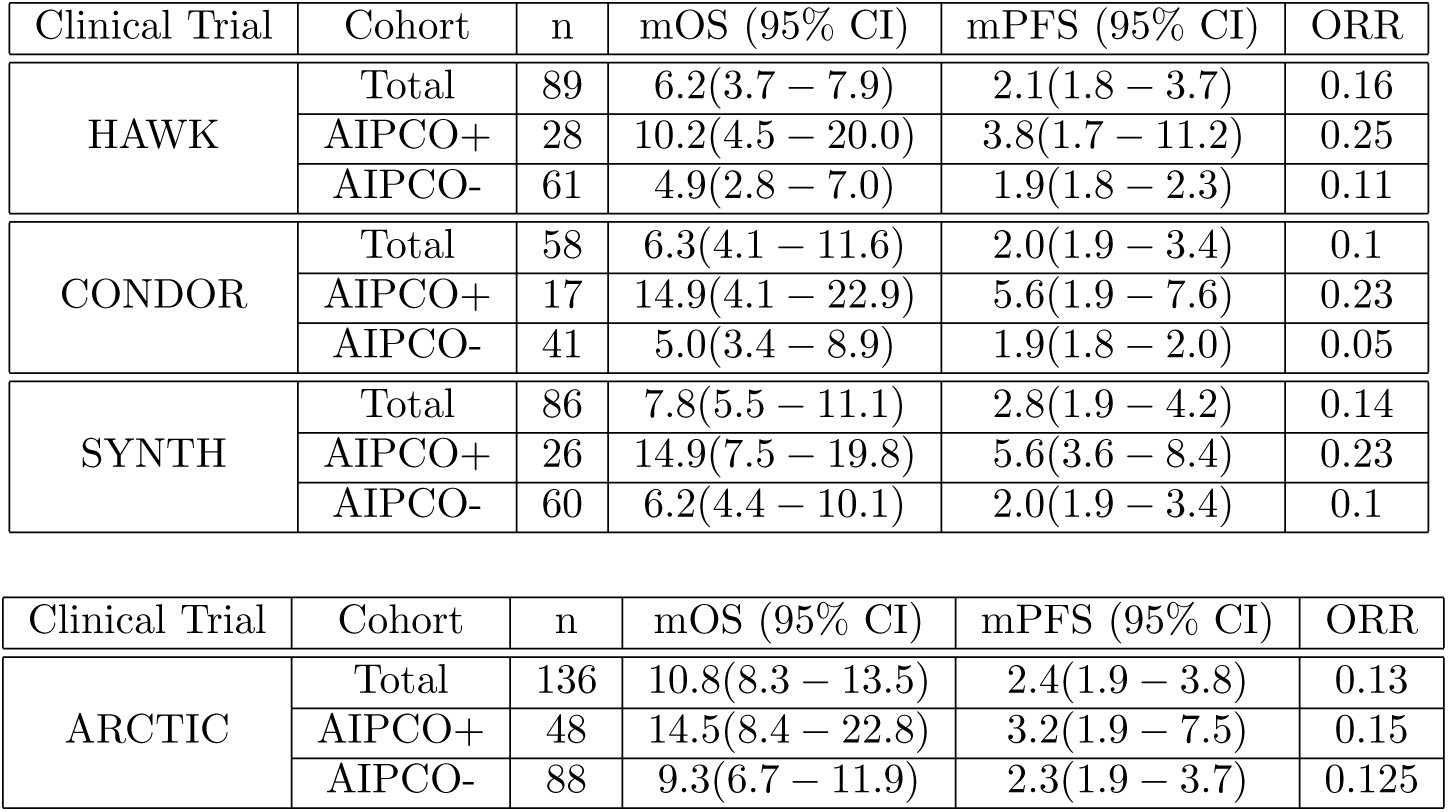
Performance of AIPCO measured in terms of median overall survival in months mOS, median progression-free survival in months mPFS, and objective response rate (ORR) for the case of head and neck squamous cell carcinoma (HAWK, CONDOR, and a synthetic cohort SYNTH composed by patients from HAWK and CONDOR), shown on the top table. The bottom table contains the performance corresponding to the non-small cell lung cancer (ARCTIC) AIPCO. In both approaches, AIPCO-positive AIPCO+ patients significantly improve for mOS, mPFS, and ORR in comparison to AIPCO-negative AIPCO-patients.

Using the Cox proportional hazards model, AIPCO+ responders in HAWK and CONDOR were associated with decreased risk of progression (HR 0.51, 95% CI 0.36-0.73 and HR 0.46, 95% CI 0.29-0.72, respectively) and as compared to the AIPCO-cohort. Overall survival followed a similar pattern as progression-free survival as AIPCO responders were associated with decreased risk of death (HR 0.58, 95% CI 0.34-0.92 for HAWK and HR 0.49, 95% CI 0.26-0.93).

The enhancemenet of efficacy of our approach is further validated using the SYNTH dataset, as illustrated in Fig. 4. The SYNTH dataset, detailed in App. A.4, comprised patient data from the HAWK and CONDOR studies to produce a dataset HNSCC with the whole range of PD-L1 status. In this case, the median OS for the AIPCO+ responders group was 14.9 months, notably higher than the 6.2 months observed for the AIPCO-group. This trend extended to the median PFS of 5.6 months for AIPCO+ patients, compared to the 2.0 months of the AIPCO-cohort. Moreover, ORR is 23% for the AIPCO+, significantly higher than 10% of the AIPCO-set. The hazard ratios for OS and PFS were 0.57 (95% CI, 0.34–0.94) and 0.50 (95% CI, 0.35–0.72), respectively, indicating a favorable risk-benefit profile for AIPCO+.

**Figure 4:**
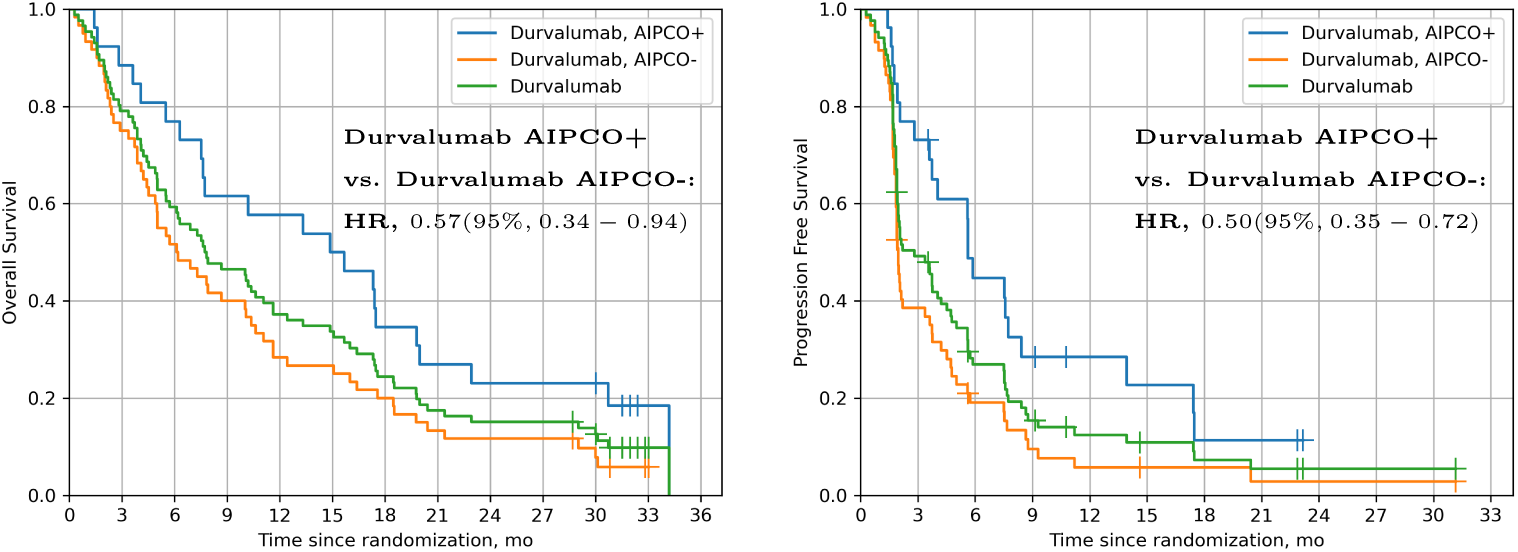
The overall survival (left) and progression-free survival (right) among SYNTH (cohort created by combining HAWK and CONDOR data) patients who received durvalumab with status AIPCO+ (blue line) vs. AIPCO-(orange line). The AIPCO stratification is based on the AIPCO algorithm trained with EAGLE data. Green lines corresponds to the curves of the SYNTH patients before stratification using AIPCO. Both figures show the hazard ratio (HR) and the 95% confidence interval (CI) for comparing AIPCO+ and AIPCO-patients.

#### AIPCO for non-small cell lung cancer

Fig. 5 depicts a clear improvement in OS and PFS for the AIPCO+ group in ARCTIC, which is numerically expressed at the bottom of Table. 1. In particular, in terms of mOS, the 48 AIPCO+ patients achieved 14.5 months compared to the 9.3 months for the 88 AIPCO-cohort. For mPFS, AIPCO+ patients reach 3.2 months, while for the AIPCO-set, 2.3 months are achieved. This improvement is also reached for ORR, with 15% of the population selected by our approach, while for the AIPCO-the calculated value is 12.5%.

**Figure 5:**
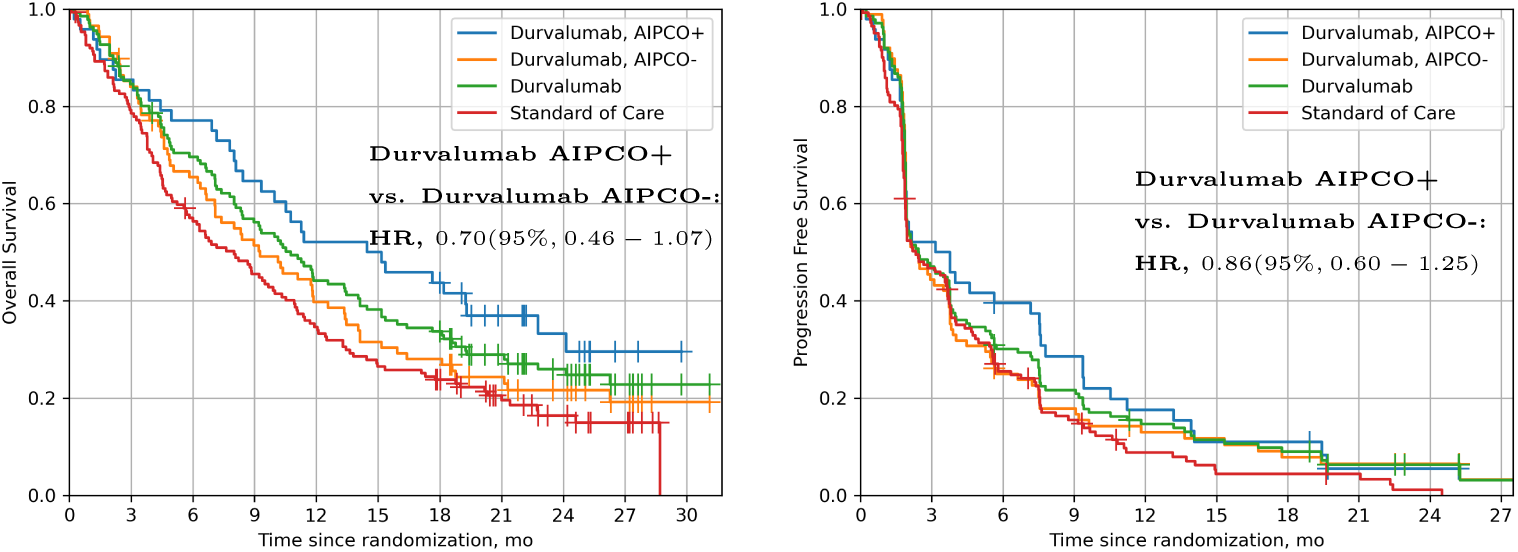
The overall survival (left) and progression-free survival (right) among ARCTIC patients who received durvalumab with status AIPCO+ (blue line) vs. AIPCO-(orange line). The AIPCO stratification is based on the AIPCO algorithm trained with MYSTIC data. Green lines correspond to the curves of the ARCTIC patients before stratification using AIPCO. Both figures show the hazard ratio (HR) and the 95% confidence interval (CI) for comparison between AIPCO+ and AIPCO-patients.

Using the Cox proportional hazards model, AIPCO+ responders in ARCTIC were associated with decreased risk of progression (HR 0.86, 95% CI 0.6-1.25) compared to the AIPCO-cohort. Overall survival followed a similar pattern as progression-free survival as AIPCO responders were associated with decreased risk of death (HR 0.70, 95% CI 0.46-1.07).

#### Comparison with PD-L1 Expression

While PD-L1 testing is performed routinely for all newly diagnosed NSCLC and HNSCC patients, clinical evidence shows that the PD-L1 biomarker neither guarantees response to first-line ICI monotherapy in patients with high PD-L1 expression nor rules out the possibility of response in patients with low or negative PD-L1 expression [46].

The AIPCO platform described in this article is a computational predictive model that analyzes pre-treatment digital pathology slides to predict clinical benefit from anti-PD-(L)1-based therapy. We found that AIPCO scores are not correlated for HNSCC (Spearman correlation coefficient *ρ* = 0.015 with p-value = 0.86) or weakly correlated for NSCLC (*ρ* = *−*0.168 and p-value = 0.051) with PD-L1 expression as determined by the VENTANA SP263 assay, as shown in Fig. 6, suggesting that the AIPCO and PD-L1 expression biomarkers are addressing discrete biological aspects of the tumor.

**Figure 6:**
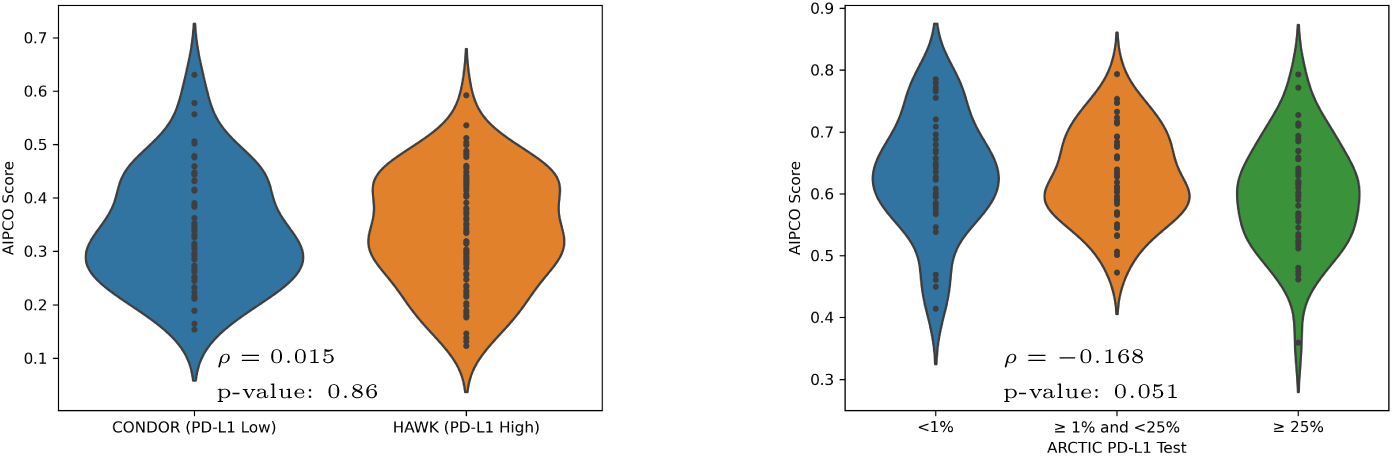
Violin plots of the AIPCO scores for head and neck squamous cell car-cinoma (left) and non-small cell lung cancer (right). For HNSCC, the Spearman correlation coefficient (*ρ*) is 0.015, with a p-value of 0.86. This analysis, comparing the AIPCO score and PD-L1 class, corresponding to CONDOR (PD-L1 Low/Negative) and HAWK (PD-L1 High), suggests that there is no statistically significant relationship between the AIPCO score and the PD-L1 status. Conversely, for NSCLC using ARCTIC data, a Spearman coefficient correlation (*ρ*) of *−*0.168 was calculated, whose corresponding p-value is = 0.051. These results may indicate a weak negative relationship between AIPCO score and PD-L1 status for NSCLC.

### 2.2 Interpretability of AIPCO model predictions

Fundamental components of AIPCO are computer vision DL models utilized to score WSIs, whose values are used to stratify patients. As described in the previous section, our proposed approach obtained meaningful and promising results as a patient stratification tool for HNSCC and NSCLC patients who received durvalumab monotherapy. However, although the training and test datasets were curated, their generation did not involve pathologist intervention. Therefore, in comparison to supervised algorithms, explaining the decision-making process followed by our algorithms becomes even more critical.

Explainability of the results enhances the trust in the solution as bugs, overfitting, and other common errors could be detected, this verification facilitates the tool adoption, but this is not the only benefit. Additionally, it has the potential to improve our understanding of disease pathomechanisms. With no human intervention, DL models allow for extracting morphological and architectural features typically not recognized by human experts. These features can be used to identify morpho-clinical correlates and to refine prognostication.

We present the first interpretable DL algorithm for H&E-based prediction of HNSCC and NSCLC response to immunotherapy. Our model robustly identified morpho-clinical correlates and could enable further prognostic refinement of patients with HNSCC and NSCLC.

We separately explore the pathological interpretations of the tumor and its microenvironment of the predicted regions for both data sets using heatmap visualizations generated employing Grad-CAM [47]. We overlaid transparent risk density maps on H&E stained WSIs, which enable pathologists to correlate the model predictions with the underlying histology of the disease. Density maps were generated using a trained AIPCO model to predict the risk for each region of interest (ROI) in a whole-slide image. The ROI predicted risks were then aggregated to a WSI level, followed by a color map to overlay on WSI, where red and blue indicate high and low AIPCO risk regions, respectively. A selection of risk heatmaps from multiple patients is presented in Fig. 7, with ROIs showing how AIPCO predictions correlated with important pathological phenomena.

**Figure 7:**
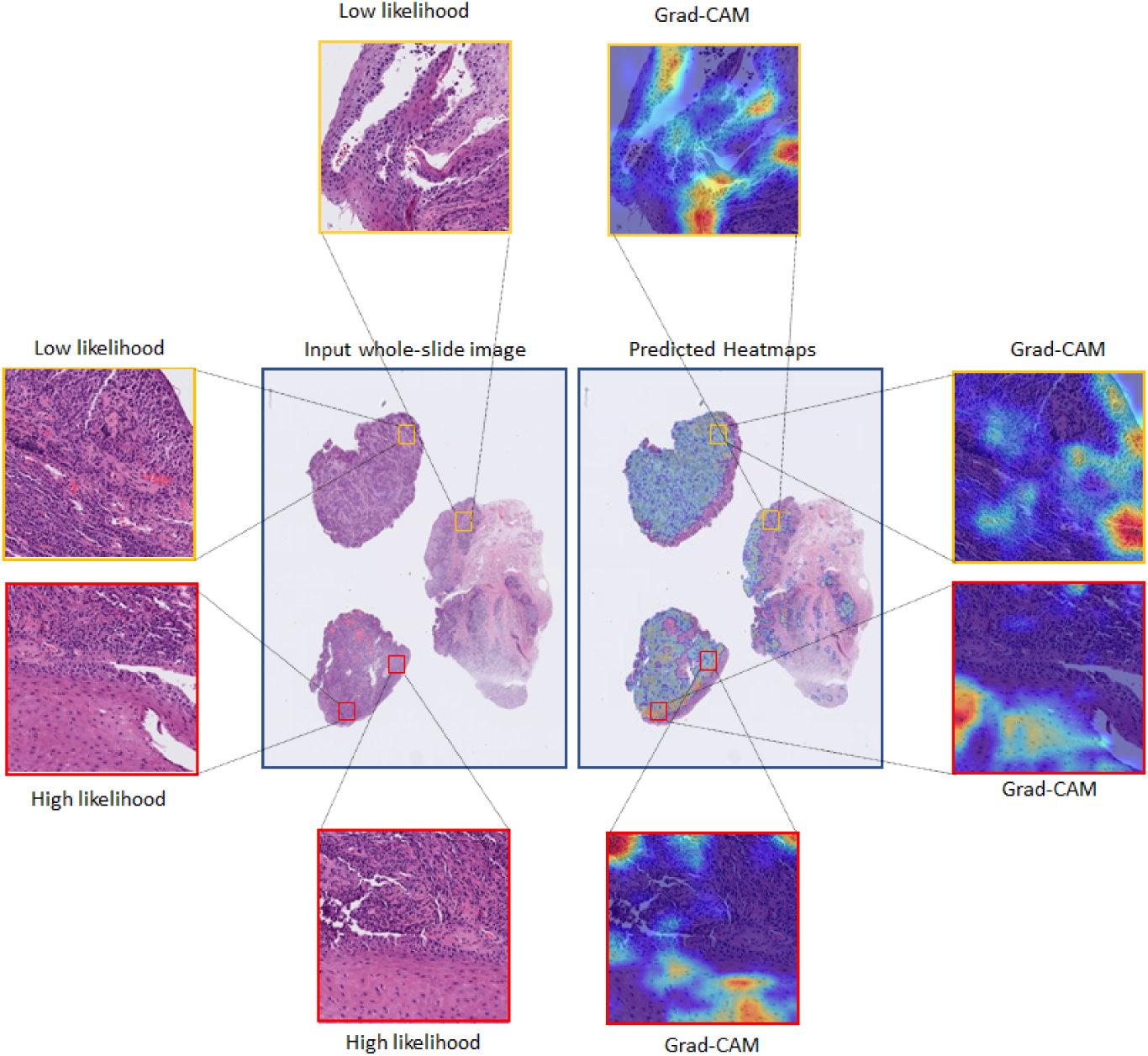
An example of the analysis of the results generated by AIPCO. On the left side, an original H&E-stained whole-slide image (WSI) with four zoomed patches corresponding to regions of the image with low likelihood score and high likelihood score. On the right part, a heatmap of the whole slide image of the AIPCO score. By zooming in, a finer analysis of the algorithm using Grad-CAM is shown at a patch level to explain which visual features were critical to assign the score to a given patch.

The AIPCO model classified image patches as a high score or a low score for predicting response to immunotherapy in HNSCC and NSCLC. The confusion matrix indicated concordant and some discordant cases between the true response to immunotherapy and the predicted image-based classification. It is very challenging to objectively assess the histology-based morphological and architectural features used by the model to correctly predict treatment response. AI-based models have the potential to detect novel tumor-immune interrelations that human experts traditionally would not focus on.

It is well known that pathologists show limited ability to identify tumors that would respond to immunotherapy when provided with minimal clinical information and especially when evaluating based on morphology alone. Despite using only histology as input for decision-making, our model can accurately predict treatment response to immunotherapy.

Quantitative evaluation of the color map overlaying the H&E slide for the high and low score images showed that the model mainly focused on the tumor region rather than the invasive border and adjacent normal tissue, as shown on the selected ROI in Fig. 8. The model seemed to show a “higher” attention towards strong nuclear atypia (e.g., Fig. 8c and Fig. 8g) than for the low score images. The high-score images also showed specific immune-related features, such as acute inflammation and a high density of tumor-infiltrating and peritumoral lymphocytes, as in Fig. 8d and Fig. 8g. Low score images highlighted high keratinizing areas Fig. 8a. In addition, some low score images displayed a more glandular architecture as in Fig. 8b and Figs. Fig. 8e-8f, whereas high-score images showed a more solid tumor growth and a low tumor-to-stroma ratio as in Figs. 8g-8h.

**Figure 8:**
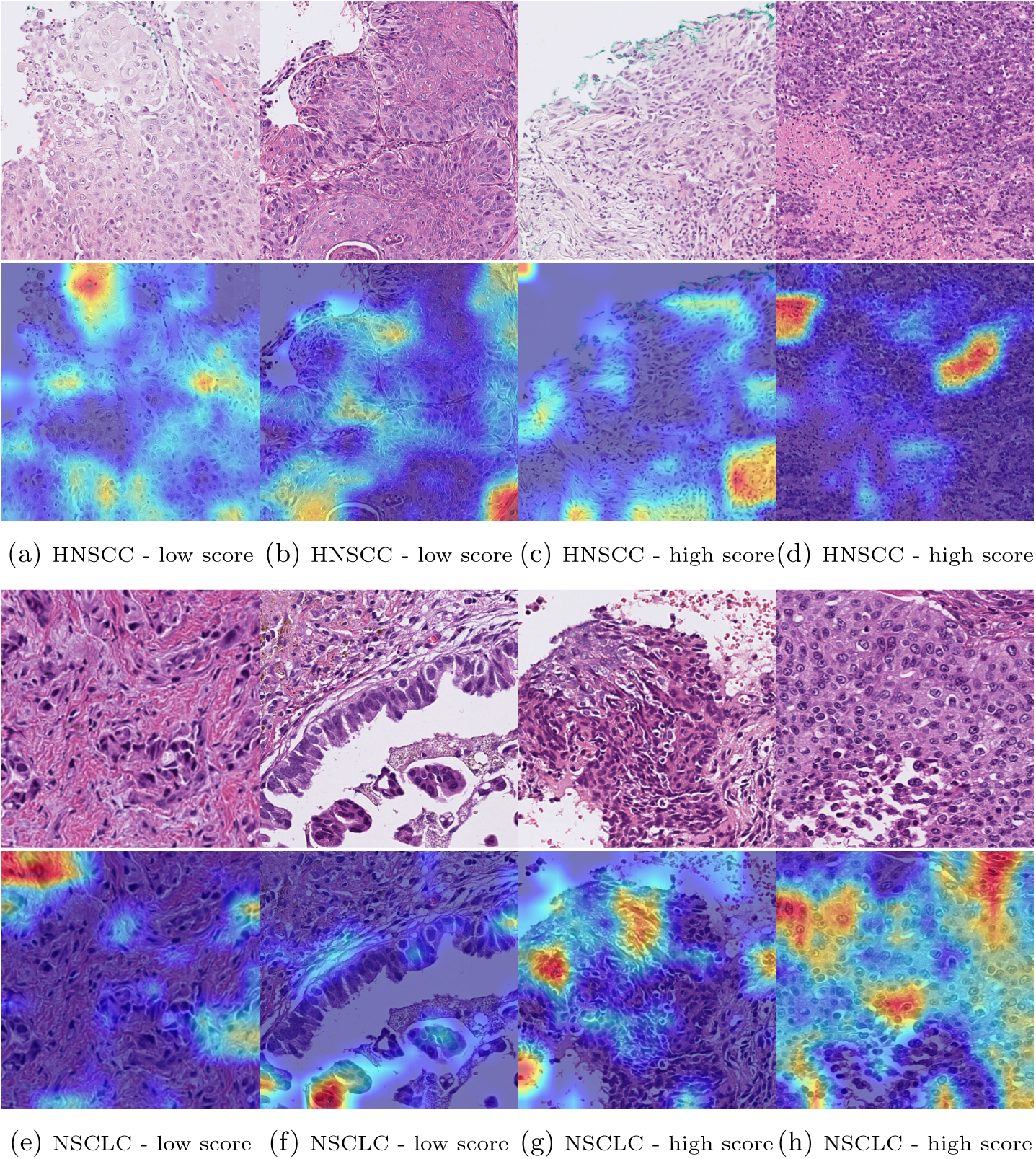
Pathological assessment and interpretability of selected AIPCO patches for head and neck squamous cell carcinoma (HNSCC) at 20*×* magnification (a-d patches), and the patches for non-small cell lung cancer (NSCLC) at 40*×* magnification (e-h images). For each of the two sets, the top row shows the selected four patches, with two low-score regions (scoring below the 25th percentile) and the other two high-score patches (scoring above the 75th percentile). In the bottom row, corresponding heatmaps generated using Grad-Cam provided insights into how decisions are made, where red corresponds to high confidence while blue is low. For HNSCC, patch (a) depicts well-differentiated characteristics and keratinization. Patch (b) is interesting, as it displays glandular and papillar patterns. The high-score patch (c) reveals nuclear atypia, while patch (d) shows clear signs of chronic inflammation. While for NSCLC, patch (e) exhibits a glandular pattern, also visible in the patch (f) with additional papillary features. Chronic inflammation with nuclear atypia is shown in the high-score patch (g), while patch (h) showcases a clear solid pattern.

## 3 Discussion

To our knowledge, this work represents the first study utilizing a machine learning approach with digitized pathology slides to predict response to an IO drug without pathologist annotations on the WSI for both HNSCC and NSCLC. Our ML algorithm, exclusively using the digitized H&E pathology slide from a pretreatment biopsy, successfully classified responders to durvalumab in a population of advanced HNSCC and NSCLC clinical trial subjects. We demonstrated that the AIPCO classification for response translated to improved OS and PFS, suggesting that this method is a clinically meaningful classification tool.

With the weak correlation of the approach to PD-L1 expression, this approach could be a reliable tool for clinicians to supplement PD-L1 expression by identifying responders to ICI based solely on digitized pretreatment pathology slides when deciding whether or not to treat an advanced cancer patient with ICI monotherapy. This is also supported for HNSCC, as AIPCO shows a statistically significant improvement for HAWK and CONDOR, which are high PD-L1 and negative/low PD-L1 studies, respectively, indicating its complementariness to PD-L1 status as a biomarker.

One of the critical questions regarding the clinical adoption of AI approaches for digital pathology in the clinic is the pathway for approval by regulatory agencies. The fundamental principle guiding the regulatory approval process is the requirement of “an explanation of how the software works” [48], and the FDA has recently started granting approval to AI-based approaches for clinical use [35]. In order to generate an AI-based companion diagnostic, developing an algorithm that shows promising performance according to the relevant metrics is only the first step. To be successful in its transition from development to implementation in the clinic, our AI approach has adopted a) good engineering practices and choosing qualified technological partners (e.g., cloud-managed services) and b) embracing good machine learning practices to mitigate the risk that algorithms will not keep the performance using real-world data.

These good practices are aligned with our focus on interpretability and explicability, which helps us identify potential overfitting issues and algorithmic inconsistencies. In particular, by analyzing how our approach made its predictions, we found features correlated with the response to IO therapy, including acute inflammation and tumor-infiltrating and peritumoral lymphocytes. On the contrary, AIPCO highlighted keratinizing areas and glandular architectures for low scores. Furthermore, by studying the visual patterns that drive the algorithm decision, we may push the science by identifying new architectural patterns in the tissue related to IO response. In parallel, running an orthogonal analysis of these cells highlighted by the algorithm with other technologies (e.g., mass cytometry or spatial transcriptomics) will allow us to characterize the biological rationale of AIPCO.

In addition, our framework shows an improvement for patients when it is trained independently for HNSCC and NSCLC, showing good generalizability properties across clinical trials. From the knowledge transferability perspective, we hypothesize the feasibility of extracting valuable features from one specific indication that could benefit another. This approach may lead to a solution that is tumor-agnostic and indication-agnostic.

Based on the promising results using retrospective data, the following steps should be running a prospective clinical trial to build confidence in this framework. In particular, we envision a clinical trial using AIPCO to prospectively select newly diagnosed recurrent or metastatic HNSCC patients to receive durvalumab monotherapy. The goal of this trial is to predict response to durvalumab based solely on a digitized pretreatment pathology slide, and as such, are targeting PD-L1 negative patients who would otherwise not be candidates for anti-PD(L)1 monotherapy. By providing prospective evidence of the validity of this AI algorithm, we will be one step closer to gaining approval for the first FDA-approved AI companion diagnostic.

Finally, from a scalability and operationalization standpoint, considering the deliberate modularity of AIPCO and its absence from pathologist annotations, we envision leveraging the full potential of machine learning operations (MLOPs) [49] to create an automated machine learning solution capable of generating candidate algorithms. On the one hand, the adoption of MLOPs would lead to the creation of an H&E-based IO-predictive algorithm factory. On the other, it would accelerate the production and the validation of new candidate solutions, for example, for new tumor types (e.g., small cell lung cancer) or new lines of therapy, including antibody-drug conjugates (ADCs). Utilizing cloud-based solutions inherently provides global reach, presenting new affordable and quick diagnostic opportunities for patients unavailable before.

## A Data-Driven Methods

### A.1 Ethical approval

The analysis presented in the manuscript uses data from patients that provided informed consent including for secondary reuse. All studies were conducted in compliance with the Declaration of Helsinki and the United States (US) Food and Drug Administration (FDA) Guidelines for Good Clinical Practice.

### A.2 Data

We used five durvalumab clinical trials to run our analysis, as illustrated in Table 2. The characteristics of the studies used for HNSCC:

**EAGLE** was a phase III study performed on recurrent and metastatic HNSCC patients to investigate the safety and efficacy of durvalumab with tremelimumab and durvalumab compared to standard chemotherapy. The primary endpoint for this study was OS, and the secondary endpoint included PFS.

**HAWK** is a single-arm multi-center phase II study of durvalumab. The patients with PD-L1 positive recurrent or metastatic HNSCC were enrolled in this study. It also included patients who progressed during or after platinum-based chemotherapy with recurrent or metastatic disease.

**CONDOR** study was a phase II, randomized, open-label study of durvalumab, Tremelimumab, and durvalumab in Combination With Tremelimumab. This study contains patients with recurrent/metastatic HNSCC.

Enrolling criteria for this study included PD-L1 low or negative disease that had progressed after platinum for both recurrent and metastatic participants. Patients were randomized (*n* = 267) from April 15, 2015, to March 16, 2016, at 127 sites in North America, Europe, and Asia Pacific. The primary outcome was to access safety, tolerability, and efficacy measured by ORR.

For the NSCLC analysis, the data utilized corresponded to:

**MYSTIC** was a phase III clinical trial study performed to investigate the safety and efficacy of durvalumab with tremelimumab and durvalumab monotherapy compared to platinum-based chemotherapy in 1st line treatment. This global multi-center study includes participants with epidermal growth factor receptor (EGFR) and anaplastic lymphoma kinase (ALK) wild-type locally advanced or metastatic NSCLC.

PFS is considered one of the significant endpoints in oncology clinical trials, and it is termed as the time from enrollment into the study (randomization) until the disease progression or death.

**ARCTIC** corresponds to the randomized phase III study containing data from multiple centers to assess the efficacy and safety of durvalumab versus standard of care (SoC) in NSCLC patients with PD-L1 positive tumors and the combination of durvalumab plus tremelimumab versus SoC in NSCLC patients with glspdl1-negative tumors in the treatment of male and female patients with locally advanced or metastatic NSCLC (Stage IIIB-IV), who have received at least two prior systemic treatment regimens including one platinum-based chemotherapy regimen for NSCLC.

**Table 2:**
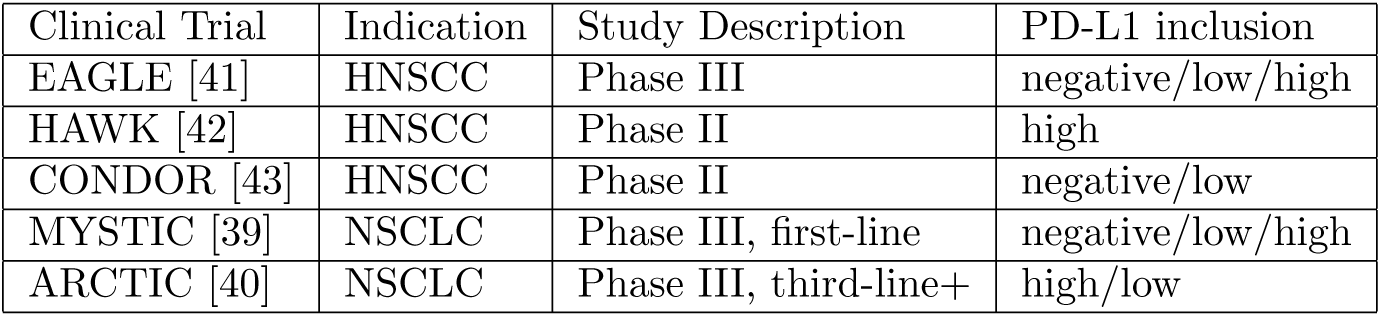
Summary of the data from five durvalumab clinical trials have been utilized to develop AIPCO. Specifically, EAGLE, CONDOR, and HAWK clinical trials for head and neck squamous cell carcinoma (HNSCC) and MYSTIC and ARCTIC were utilized for non-small cell lung cancer (NSCLC). The table includes the clinical trial phase and the PD-L1 status of the inclusion criteria.

| Clinical Trial | Indication | Study Description | PD-L1 inclusion |
| --- | --- | --- | --- |
| EAGLE [41] | HNSCC | Phase III | negative/low/high |
| HAWK [42] | HNSCC | Phase II | high |
| CONDOR [43] | HNSCC | Phase II | negative/low |
| MYSTIC [39] | NSCLC | Phase III, first-line | negative/low/high |
| ARCTIC [40] | NSCLC | Phase III, third-line+ | high/low |

### A.3 Hardware and software

All experiments were conducted on AZ’s Scientific Computing Platform using NVIDIA Tesla K-80 GPUs with 12 GB of internal memory. We program in Python (version 3.6.10), PyTorch (version 1.31.), and Torchvision (version 0.4.2) as the deep learning framework. Some key packages used were OpenSlide (version 1.1.1) to process and analyze the WSIs, and for the general data analysis, we used Numpy(version 1.19.1), Pandas (version 1.1.0), and Lifelines (version 0.25.8) for the survival analysis.

All the WSIs were scanned using the Hamamatsu C13220 with a maximum magnification of 40*×*, corresponding to approximately 0.23 microns per pixel. Each WSI is represented using a pyramid format that contains the image represented with different magnifications, including 20*×*, corresponding to 0.46 microns per pixel and 0.92 microns per pixel for 10*×*.

### A.4 Datasets for training, cut-off selection, and testing

In data-driven algorithm development, the data is usually employed for training, validation, and testing, where in order to show generalizability, the train and the test set must be independent. The test set comprises data from patients who have not been part of the information during the training process. Following this methodology, we assess how effectively our solution can handle unseen information.

To enhance the applicability of our approach in the real-world setting, we have implemented an additional condition for the test set. This requirement ensures that the test set consists only of data from clinical trials not used during training. Consequently, the results reported not only take into account the variations among patients but also consider the variability produced by different clinical trials. This strategy provides more reliable information about the algorithm’s readiness as a patient stratifying tool. This is aligned with the best machine learning practices and the guidelines created by the FDA [50].

In the case of training a drug response prediction for HNSCC and NSCLC, as shown in Table 3, we divided the available AZ’s durvalumab datasets into three categories according to their use:

- The data corresponding to patients of the durvalumab arm from EAGLE or MYSTIC are used for training purposes for the HNSCC and NSCLC, respectively.
- Information from patients of the SoC arm from EAGLE or MYSTIC is used to determine the cut-off for stratification.
- With all AIPCO model parameters defined, in the case of the HNSCC, the data corresponding to patients from HAWK and the durvalumab arm from CONDOR are utilized for testing. In the case of NSCLC, ARCTIC patients who received durvalumab were used for testing.

**Table 3:** Immunotherapy clinical trials and data used for developing and testing the AIPCO algorithms for head and neck squamous cell carcinoma (HNSCC) (top table) and non-small cell lung cancer (NSCLC) (bottom table). Both for the HNSCC and NSCLC development, the durvalumab monotherapy arm and standard of care (SoC) arms were used for training and cut-off determination, respectively. For testing, HAWK and CONDOR datasets and the synthetic cohort SYNTH were used for HNSCC, while ARCTIC durvalumab monotherapy patients’ data were in the case of NSCLC. In both tables, it is shown the number of whole-slide images original part of the dataset, as well as the number of slides after curation.

| Clinical Trial | Arm | Purpose | # WSIs | # curated WSIs |
| --- | --- | --- | --- | --- |
| EAGLE | Durvalumab | Training | 123 | 122 |
| EAGLE | SoC | cut-off selection | 122 | 122 |
| HAWK | Durvalumab | Testing | 109 | 91 |
| CONDOR | Durvalumab | Testing | 60 | 60 |
| SYNTH | Durvalumab | Testing | 94 | 88 |
| MYSTIC | Durvalumab | Training | 312 | 296 |
| MYSTIC | SoC | cut-off selection | 297 | 271 |
| ARCTIC | Durvalumab | Testing | 146 | 144 |

In the case of the algorithm developed for HNSCC, we can test the efficacy of the solution for PD-L1 positive patients using HAWK data and for PD-L1 low or negative patients using CONDOR data. Complementary to this, it is worth evaluating the performance characteristics of the algorithm for the scenario of all-comers. To achieve this, we create a new cohort, denoted by SYNTH in this manuscript, by combining patients from both HAWK and CONDOR studies so that its PD-L1 distribution closely resembles the expected PD-L1 score distribution for all-comers. We approximate the probability of PD-L1 scores for all-comers as P(PD-L1 *<* 1%) = P(PD-L1 *≥* 1% *∪*PD-L1 *<* 50%) = P(PD-L1 *≥* 50%) = 1*/*3, where P(*·*) denotes the probability that a patient’s PD-L1 score falls within the specified interval. As the PD-L1 cut-offs provided in the available clinical information are not concordant with this desired distribution for all-comers, we combined HAWK and CONDOR datasets by pseudo-randomly selecting patients with the following distribution:

- 48 patients from HAWK with a PD-L1 score greater than 25%,
- 15 CONDOR patients with PD-L1 scores ranging from 1% to 25%,
- and 31 patients from CONDOR with PD-L1 score below 1%.

The cohort of ARCTIC durvalumab patients used in this work is obtained as the combination of the patients of the ARCTIC Sub-study A, whose patients show PD-L1 status high during the pre-screening period and patients from Substudy B that were classified as PD-L1 low.

### A.5 Whole slide image sampling

WSIs usually have gigapixel resolution, producing a challenge for applying outof-the-box computer DL approaches due to limitations in the typical memory size of GPUs. For example, the minimum size of a batch of complete WSIs exceeds the memory capacity, making these approaches infeasible.

To address this limitation in the adoption of DL techniques, we implemented a common approach of sampling the WSI, generating smaller patches that are suitable for use in GPUs. For each *i*-th patient, its corresponding WSI was sampled using a tile of the size of 512 *×* 512 pixels for the magnification selected for the algorithm (e.g., 40*×*, 20*×*, and 10*×*).

The patches were produced by pseudo-randomly uniformly distributed sampling within the area of the digital pathology image. Per WSI, *N* tissue patches are generated, focusing on the area of the image that contains tissue, dismissing the area of the WSI containing the background. This process removes data that is not relevant for the algorithm development, reducing the computational needs. Fig. 9 shows a visual example of the outcome sampling process with a 10*×* magnification of a WSI corresponding to a CONDOR patient. We use **w***^i^* to denote the *j*-th tissue patch of the *i*-th patient and *W^i^*for the corresponding set of patches, where *j* = 1*, …, N*.

**Figure 9:**
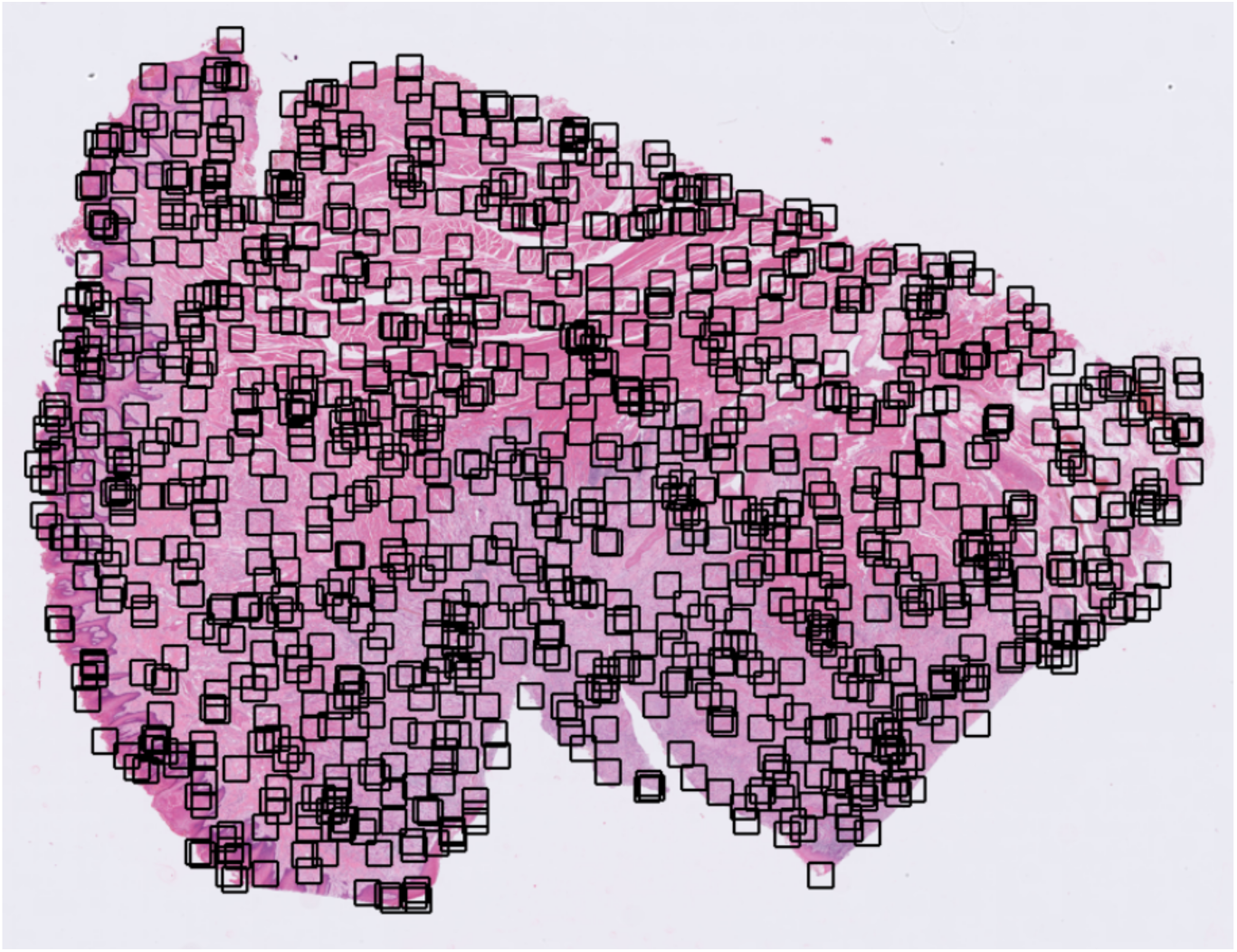
A visual example of a hematoxylin and eosin-stained whole-slide image corresponding to a head and neck squamous cell carcinoma patient. The image shows, as black squares, the pseudorandomly-obtained 10^3^ 512 *×* 512 pixels patches generated at a 10*×* magnification.

### A.6 Cancer whole-slide image patch selection

We hypothesize that the most relevant information needed to discriminate between IO responders can be obtained by analyzing the tumor microenvironment. As a part of the algorithm pipeline, we implement this by classifying the WSI tissue patches into two classes: those that contain tumor cells and those that do not. This step in the context of the whole algorithm is shown on the top row in Fig. 1.

In particular, given the *i*-th patient, the corresponding tumor patches obtained from WSI *W^i^* are classified using a cancer cell classification DL model. Then, the patches classified as positive create a new set, *W∼^i^*, that are the images used by our framework either for training or testing purposes.

In addition, by using this tumor patch classifier, our approach removes patches from the algorithm that could hinder its performance. For example, this process could eliminate the patches that show quality issues, including margin ink, bubbles, dark spots, tissue folding, pen marks, fingerprints, and out-of-focus. By running this step, we reduce the overall risk related to the reliability or generalizability of the produced algorithm.

Fig. 10 depicts a WSI heatmap visualization of the cancer cell patch classifier corresponding to a CONDOR patient using a sliding window to produce the normalized score. The details of the creation of the tumor patch classifier can be found in App. A.6.1

**Figure 10:**
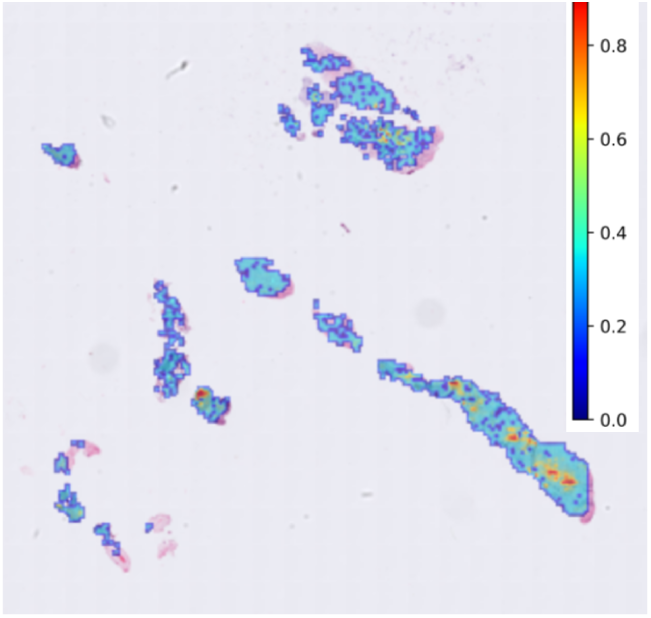
Example of a whole-slide image (WSI) heatmap of a cancer cell patch classifier score of a CONDOR patient. This score helps to define the regions of interest of WSI that our algorithm uses to decide between the patients who respond to durvalumab and those who do not.

Table 3 illustrates how the number of WSIs available for development is reduced by the curation process that is used to enhance the quality of the data. WSIs are dismissed if they do not contain any cell patch classified as cancerous. As a result, 122 EAGLE durvalumab monotherapy WSIs are used for training the HNSCC model, and 296 curated MYSTIC WSIs are employed for the same purpose for the NSCLC case.

#### A.6.1 Cancer patch classifier model development

As shown in Fig. 1, one of the components of our framework is the cancer patch classifier. This model is employed to pinpoint the regions within WSIs with tumor tissue as the areas containing the most relevant information about the disease, including its nature and response to treatments.

This DL model was developed using the PatchCamelyon (PCam) data [32, 51]. This dataset has 327680 96 *×* 96 color 10*×*-magnified images. The labels of the dataset indicate if the patch contains tumor tissue. For generating this model, we modified the dataset defined for the Histopathologic Cancer Detection Kaggle competition [52]. Specifically, we divided the original training set, which comprises 220025 patches, into training and testing for our cancer patch classifier development: 80% of the patches were 176020 for training, and the remaining 20% corresponded to 44005 images for testing.

We trained an ImageNet-pre-trained VGG16 CNN [53], where the last fullyconnected (FC) layer was substituted by a multilayer perceptron composed of an FC layer with 4096 inputs and 256 outputs connected to a ReLU layer followed by an FC layer obtaining two output values that are processed through a logarithmic softmax activation function. During training, all the original layers are frozen, and only the new two FC layers are updated during training.

We used cross-entropy as the cost function for training during 25 epochs in batches of 64 patches. As the optimizing algorithm, we employed the stochastic gradient descent algorithm with a learning rate of 10*^−^*^3^ and 0.9 as the momentum. We synthetically increased the number of samples to mitigate the risk of overfitting by performing data augmentation. In particular, we pseudorandomly obtained resized crops with scale in the range [0.08, 1.0] and aspect ratio within [3*/*5, 4*/*3] before resizing. In addition, each modified patch was horizontally flipped with a probability of 0.5

The final model selected as our cancer patch classifier was the one that maximizes the accuracy in the testing stage calculated after each training epoch. In this case, the best-performing model was obtained after 11 epochs, showing an accuracy of 89.44%, whose threshold used to differentiate between positives and negatives is 0.5.

### A.7 Outcome prediction training

#### A.7.1 Weakly-supervised labeling

Given the low cardinality of the datasets used, the problem is formulated as a *k*-class classification problem. Training and test data 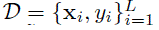, where **x***_i_* denotes the *i*-th curated patch of the set 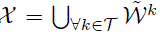, where *T* represents the set of patient indices belonging to the training set. *y_i_* denotes the *i*-th true label, and *L* stands for the cardinality of the training set.

In order to train the models, each patch of a given subject has an associated label corresponding to the target of the WSI. The DL model is trained to map this correspondence. The label is obtained as a function of the clinical data of the patient. Examples of the outcome of the clinical data are objective response (OR), PFS, and OS. For example, in the case of OR,

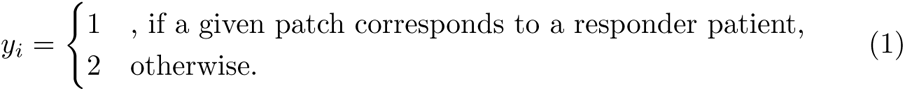

Or, in the case of using OS, one can use a threshold T (e.g., T = 18 months) to formulate the task as a binary classification problem as

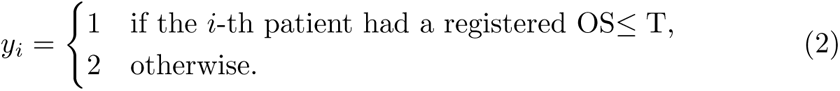

Given the *i*-th patient, the label is assigned to all the patches extracted from the WSI. However, it is sensible to think that not all the patches containing cancer cells are accurately associated with the clinical outcome. Therefore, one should address the problem as a noisy supervised task or weakly supervised machine learning approach.

#### A.7.2 Deep ensemble for predicting clinical outcome

Developing a solution to predict clinical outcomes for IO therapy only using H&E-stained slides presents its challenges, requiring tailored approaches. For example, due to the limited number of patient samples, the available data for algorithm training and testing is restricted.

In this low-data regime scenario, it is well-known that a classic one-model solution often shows a tendency to overfit, lack of robustness, and consequently bias. Therefore, we needed to focus on ML techniques that are efficient in using data.

Furthermore, the complexity of the challenge significantly increased as the disease pathomechanisms that drive the response to IO are incompletely understood, and the WSIs are not annotated. Therefore, we addressed our goal by formulating it as hypothesis-free and weakly supervised.

Consequently, we designed and developed a solution to mitigate these risks by combining transfer learning and deep learning ensembles. Transfer learning enables the transfer of knowledge learned from a different task to a related task, thus reducing the demand for data, while deep learning ensembles combine several individual models to generate more robust and generalizable algorithms. Ensemble learning, as demonstrated by random forest [54] or XGBoost [55], consistently performs well across various applications.

One of the main issues regarding the use of DL ensembles is their high computational requirements. For instance, an ensemble composed of *M* models run sequentially would increase the execution time by a factor of *M*. In addition, there are complexities regarding maintaining *M* models during their entire lifecycle compared to one model. Nevertheless, more importantly, interpretability and explainability are critical to facilitate the adoption of hypothesis-free approaches like ours. This would include the independent analysis of *M* DL algorithms and the fusion of their corresponding scores that become time-consuming and complex tasks. To address these issues, we incorporated knowledge distillation [56–58] in our framework. Knowledge distillation is the process of transferring knowledge from a larger trained model to a smaller model and has shown promise in digital pathology applications [59, 60].

In our framework, the ensemble of *M* DL models acts as a teacher for a new CNN, which is trained to replicate the teacher’s outcomes per dataset. Knowledge distillation helps to resolve scoring discrepancies among the baseline models and serves as a regularization tool that improves the robustness of our solution. In this way, the student CNN will potentially perform as the teacher while the computational cost and maintenance effort are significantly reduced. Moreover, the interpretability and explainability become more straightforward, as there is only one model to analyze.

In summary, our framework for developing algorithms to predict the clinical outcome of immunotherapy using unannotated H&E WSIs comprises three stages, as illustrated in Fig. 11. In Stage 1, *M* deep learning baseline models are trained. In Stage 2, *M∼* models are selected. Finally, in Stage 3, knowledge distillation transforms the ensemble of original *M* baseline models into a single DL model.

**Figure 11:**
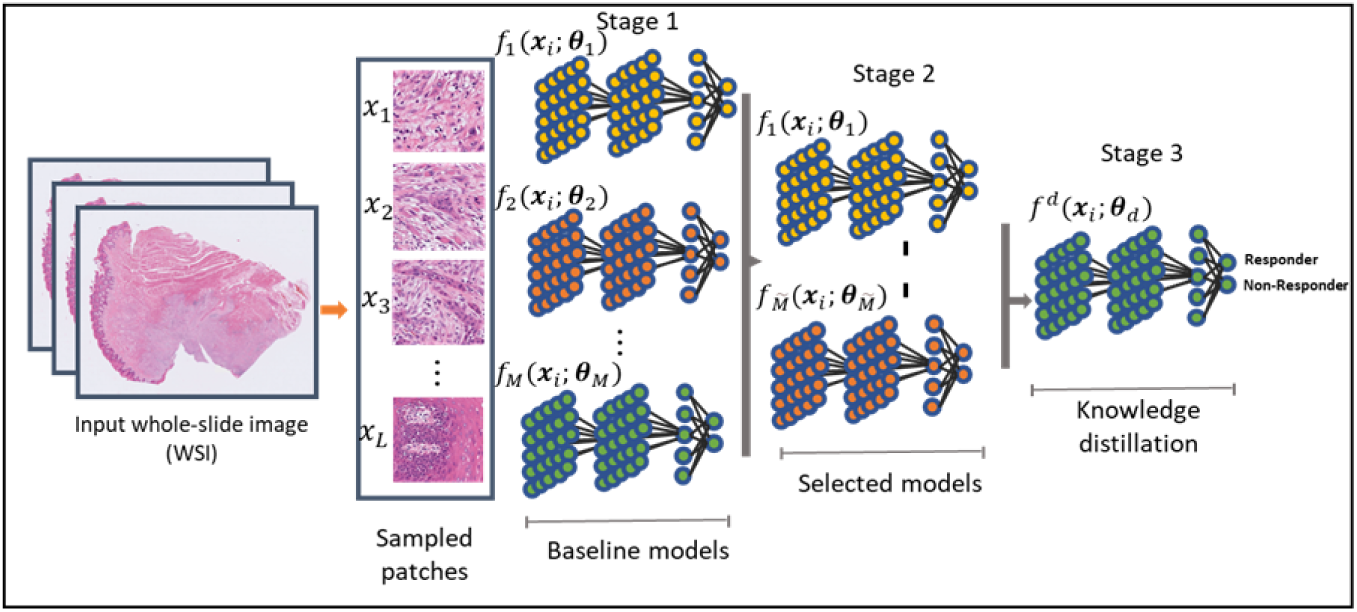
Overview of the process to obtain the AIPCO model based on ensemble learning and knowledge distillation. First, the sampled tumorpositive patches **x***_i_*obtained from the H&E from the training set of the whole-slide image, where *i* = 1*, …, L*. Then, in Stage 1, *M* baseline models 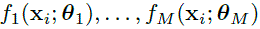 are trained with respect to the clinical outcome (e.g., objective response or overall survival). In Stage 2, *M∼* models are selected based on the performance of the validation set from the *M* trained models. Finally, at Stage 3, the final model is computed using knowledge distillation of the assemble of the output of the selected *M∼* models to obtain one single final AIPCO model 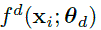 to stratify patients into two categories Responder AIPCO+ and Non-Responders AIPCO-.

### Stage 1: Generation of the baseline models

We chose to obtain a variety of baseline models for the ensemble by using different subsets of the training set. In particular, to capture the sample variations, we pseudorandomly split *M* times the training set (i.e., EAGLE and MYSTIC durvalumab arms for HNSCC and NSCLC, respectively) into baseline training set with approximately 80% of the patients and 20% of them for the validating set of the baseline models.

We trained the algorithm for each of the *M* splits using the baseline training set, evaluating the baseline models with their corresponding baseline validation split. This enables a coarse exploration of the feasibility of capturing the clinical outcome using a particular hyperparameter configuration (e.g., backbone, magnification, optimization technique, etc.). For example, for the curated *i*-th patch **x***_i_*, the corresponding score 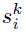 is computed by the *k*-th baseline model 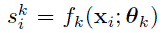, with *k* = 1*, …, M* and ***θ****_k_* representing the parameters of the *k*-th baseline model.

As the backbone for the baseline models, we employed ResNet50 [61]pre-trained on ImageNet. We substituted the last FC layer with a stack composed of one FC layer with 2048 inputs and outputs, followed by a Rectified Linear Unit (ReLU) connected to a second FC layer with 2048 inputs and two outputs. Finally, during training, we used a logarithmic softmax layer as the last layer. All the layers are frozen except the ones corresponding to the stack added, as this helps minimize the number of trainable parameters, thus reducing the risk of overfitting. Stochastic gradient descent (SGD) is utilized to minimize the cross-entropy loss function in batches of 32 patches of WSIs. The optimization process uses a momentum of 0.9 and a weight decay of 5 *×*10*^−^*^5^ over 100 epochs. The learning rate is set to 10*^−^*^3^ for the first 80 epochs and subsequently reduced to 10*^−^*^4^ for the remaining 20.

Moreover, it is common for the baseline training sets to show class imbalance; however, in our application, it is crucial that the produced models are not biased toward any specific category. To mitigate potential drift during training, we balanced the training datasets by oversampling the categories with fewer samples.

As mentioned, given the computational constraints of the current GPUs, the WSIs must be sampled to facilitate the training. Once the DL models are trained, it is necessary to fuse the patch-level scores at a patient level to evaluate the discriminative capability between responders and non-responders. We evaluated several approaches for aggregating the results, including taking the mean, the median, and the ratio between the number of patches, taking a value larger than or equal to a certain value, and the total number of patches. Based on the results, we decided to prioritize the mean of the patch scores, as it provided good characteristics in terms of performance, robustness, and explainability. Therefore, for a given *N∼*-length array of patch scores corresponding to the *i*-th patient 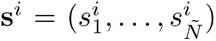, the aggregated patient-wise score *s^i^* is calculated as the arithmetic mean 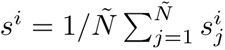.

### Stage 2: Model selection

By design, our approach creates several baseline models, each showing their unique performance characteristics as a consequence of using different baseline training sets (e.g., using different subsets of the training set). To produce an accurate ensemble, it is necessary to differentiate between baseline models that enhance the average performance of the ensemble for a given criterion (e.g., median OS or median PFS) and those that deteriorate it. In this manner, by only using the outputs of the algorithms that produce more accurate decisions, the overall performance of the ensemble will be inherently improved.

We defined the criterion for selecting baseline models identifying positive patients (e.g., responders according to RECIST 1.1. criterion), while false positives are zero. This criterion can be algorithmically implemented by analyzing the precision-recall curves and selecting the *M∼* baseline models with a precision of one for recall values larger than zero.

### Stage 3: Knowledge distillation

The knowledge of the selected *M∼* models is distilled to produce one final model, which is achieved in two steps:

- In the first step, each of the *M∼* selected models produces the scores of the curated patches corresponding to its baseline test set. As a patient can appear in the baseline test set of several baseline models, the distilled score is calculated as the maximum of the scores per patch for each WSI to prioritize high-score patches.
- In the second step, a new CNN is trained using the distilled scores of the curated patches using the Kullback-Leibler divergence (KLD) as the loss cost function. The KLD loss enables the student to learn a similar distribution of information as the teacher, thus optimizing the use of the information. This model is trained using the same hyperparameters as the baseline models but only for 20 epochs.

After the knowledge-distilled model is obtained, the cut-off needs to be computed to map the continuous score into two classes: AIPCO+ for patients who are likely to benefit from IO therapy and AIPCO-for those who would not. In order to do this, we employed the EAGLE/MYSTIC SoC arms, in which the patients were not treated with immunotherapy; however, the distribution of the scores should be consistent since the tissue was obtained in the same manner as in the durvalumab arms. We selected the 75th percentile of the patients’ scores in those arms as the cut-off for our algorithms. With the model trained and the cut-off established, the final AIPCO model can be used in a different dataset to validate its generalizability.

#### A.7.3 Generation of the head and neck squamous cell carcinoma and non-small cell lung cancer models

We employed the methodology proposed in the previous section to develop and lock all the parameters of the HNSCC and NSCLC algorithms. In order to make the algorithm easy to maintain, it is crucial to keep simplicity in the algorithm design. Aligned with this principle, we developed an algorithm using only one magnification. Through preliminary simulations, we decided that a magnification of 10*×* was appropriate for HNSCC, while 20*×* was chosen for NSCLC.

To demonstrate the versatility of our approach, we focused on training the baseline models using objective response for HNSCC. Conversely, for NSCLC, we trained deep learning models using patches derived from patients with a survival duration exceeding 18 months (Overall Survival ¿ 18 months).

Respectively for HNSCC and NSCLC, Tables 4 and 5 contain the median OS as the sequential steps of the algorithm are computed to finally generate the distilled model. The data shown for the case of the baseline ensemble and selected ensemble are obtained based on the union of the set of AIPCO+ patients of the models, where the patients belong to the test set of each baseline model. Therefore, these results correspond to samples of patients that were not used in the deep learning training of the solution. As for the distilled model, the data used for knowledge distillation is generated as a function of the scores of the test set; therefore, the analysis of data must take this into consideration.

**Table 4:** Median overall survival (mOS) in months and the 95% confidence interval for the *n* patients per cohort receiving durvalumab during the EAGLE clinical trial for head and neck squamous cell carcinoma (HNSCC). The results were obtained using stratification of our development framework at each stage to classify patients between AIPCO+ and AIPCO-. Complementary to this, the results of the total number of patients per stage without employing our framework are displayed. In stage 1, an ensemble of the *M* = 20 baseline models is utilized. In Stage 2, the ensemble was refined using the selected *M∼* = 9 baseline models. Finally, Stage 3 produces the knowledge-distilled solution.

| EAGLE | Stage 1, $M = 20$ | | Stage 2, $\tilde{M} = 9$ | | Stage 3 | |
| --- | --- | --- | --- | --- | --- | --- |
|  | n | mOS (95% CI) | n | mOS (95% CI) | n | mOS (95% CI) |
| Total | 119 | 7.6(5.2 – 9.9) | 103 | 6.3(4.7 – 8.1) | 103 | 6.3(4.7 – 8.1) |
| AIPCO+ | 30 | 8.13(5.1 – 22.6) | 25 | 14.7(5.1 – 22.6) | 25 | 10.2(4.5 – 22.6) |
| AIPCO- | 89 | 7.3(4.67 – 9.8) | 78 | 5.5(4.0 – 7.7) | 78 | 6.1(4.0 – 8.1) |

**Table 5:** Median overall survival (mOS) in months and the 95% confidence interval for the *n* patients per cohort receiving durvalumab during the MYSTIC clinical trial for non-small cell lung cancer (NSCLC). The results were obtained by using stratification of our development framework at each stage to classify patients between AIPCO+ and AIPCO-. Complementary to this, the results of the total number of patients per stage without employing our framework are displayed. In stage 1, an ensemble of the *M* = 20 baseline models is utilized. In Stage 2, the ensemble was refined using the selected *M∼* = 11 baseline models. Finally, Stage 3 produces the knowledge-distilled solution.

| MYSTIC | Stage 1, $M = 20$ | | Stage 2, $\tilde{M} = 11$ | | Stage 3 | |
| --- | --- | --- | --- | --- | --- | --- |
|  | n | mOS (95% CI) | n | mOS (95% CI) | n | mOS (95% CI) |
| Total | 268 | 14.3(10.7 – 16.6) | 252 | 14.3(10.7 – 16.6) | 252 | 14.3(10.7 – 16.6) |
| AIPCO+ | 36 | 19.2(9.3 – ∞) | 33 | 22.8(9.8 – ∞) | 20 | 23.6(8.7 – ∞) |
| AIPCO- | 232 | 13.7(10.4 – 16.3) | 219 | 13.7(10.1 – 16.3) | 232 | 13.8(10.2 – 16.4) |

The proposed framework to develop one model to predict clinical outcome per indication starts by training a set of baseline models with *M* = 20. From these candidate baseline models, a subset is selected by the criterion expressed above; for the case of HNSCC, the models meeting the criteria is *M∼* = 9, and for the case of NSCLC, *M∼* = 11. In both cases, by comparing the baseline ensemble vs. the ensemble of the selected algorithms, one can see that there is a significant improvement in the performance. In particular, for the case of HNSCC 30, AIPCO+ patients with mOS 8.13 months become 25 AIPCO+ patients with mOS 14.7 months. Analogously for NSCLC, 36 AIPCO+ patients with mOS 19.2 months are transformed into 33 AIPCO+ patients with mOS 22.8 months due to the model selection as part of our framework. Therefore, these results validate the hypothesis that by building an ensemble of models that are precise, the resulting solution improves.

This favorable trend is confirmed when one also compares the obtained data between the baseline ensemble and the final model obtained as a result of carrying out the knowledge distillation of the ensemble of the selected models. In particular, for the case of HNSCC, the baseline ensemble shows a median OS of the AIPCO+ patients of 8.13, while in the distilled model becomes 10.2 months. Similarly, in NSCLC, the cohort of AIPCO+ patients of the baseline ensemble model obtains a mOS of 19.2 months, while in the knowledge-distilled model, the resulting median OS is 23.6 months.

Based on the results, these models, one for HNSCC and another for NSCLC, support the use of the knowledge distillation process for both indications. The data presented in Sect. 2 was obtained with them, supporting its generalization across datasets.

## B Performance metrics

### B.1 Overall survival

The overall survival (OS) is generally regarded as the primary endpoint to evaluate the patient’s response to treatment for most of the oncology clinical trials. OS is defined as the duration from the date of diagnosis or start of treatment to the time of the event.

### B.2 Progression-free survival

The progression-free survival (PFS) is a clinical endpoint in clinical trials assessing treatment efficacy, especially in scenarios where OS takes a long to time to observe. It measures the time between the treatment starts and the occurrence of disease progression or other events, including death.

### B.3 Objective response rate

The objective response rate (ORR) is used in routine clinical practice and is a parameter to demonstrate the effectiveness of an oncology-related treatment. It represents the proportion of patients with a reduction in disease burden of a predefined amount.

### B.4 Precision and recall

Precision is a metric to evaluate the positive predictive ability of a classifier and is defined as the ratio between correctly classified positive incidents and the total number of positively classified incidents (true positive or true negative). Recall, on the other hand, is a metric to evaluate the ability of a classifier to correctly identify all incidents of a relevant class and is defined as the ratio between the correct predictions from the selected class and the total number of samples from the class. Mathematically, this is expressed as:

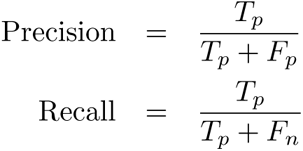

where *T_p_*, *F_n_*, and *F_p_* represent the number of true positive, false negative and false positive incidents. Given a validation dataset, each correctly identified prediction is classified as either true positives or true negatives, whereas misclassified predictions are categorized as false negatives and false positives.

## Data Availability

All data produced in the present study are available upon reasonable request to the authors

## Acknowledgements

We express our deepest gratitude to AstraZeneca’s Scientific Computing Platform, AI Bench, Imaging Platform, and Data Provisioning Operations for their critical support. Special thanks to Marietta Scott and Michel Vandenberghe for their work in the initial stages of this study. We thank Nicola Lawson for her contribution to digitizing pathology slides for this study. We are thankful for the enriching discussions with Phil Jewsbury, Marlene Dressman, and Ross Stewart, and for the valuable insights on the clinical application of AI from Leora Horn and Lee Krug. Gratitude is also extended to Irina Knyazeva for her help in the strategic vision, to Renee Iacona for Oncology Biometrics support, and to Hisani Madison, Janaki Veeraraghavan, Kelly Oliner, and Alexander Kohlmann for their contributions to our understanding of AI in diagnostics. We also thank Jason Hipp for his expertise in AI-based digital pathology during the initial phase of our work. Finally, we extend our appreciation to Ruth March, whose support and great leadership were pivotal to the success of this study.

